# Characterizing Stroke Clots Using Single-Cell Sequencing

**DOI:** 10.1101/2025.02.06.25321828

**Authors:** Daniela Renedo, Tanyeri Barak, Jonathan DeLong, Julian N. Acosta, Nanthiya Sujijantarat, Andrew Koo, Joseph Antonios, Cyprien Rivier, Santiago Clocchiatti-Tuozzo, Shufan Huo, James Giles, Guido J Falcone, Kevin N Sheth, Ryan Hebert, Murat Gunel, Lauren H Sansing, Dhasakumar S Navaratnam, Charles Matouk

## Abstract

**Background:** Ischemic stroke result in significant morbidity and mortality. By examining gene expression of cells comprising stroke clots, we aim to gain valuable insights into the underlying mechanisms of this disease and identify potential biomarkers of stroke etiology.

**Methods:** We employed single-cell RNA sequencing to analyze 10 clot samples from patients diagnosed with large vessel occlusion stroke. We aimed to identify and compare the immune cell compositions and gene expression profiles between stroke clots (atrial fibrillation vs carotid atherosclerosis). We also used MAGMA and GWAS summary statistics from the GIGASTROKE consortium to assess associations association between genetic variants and cell type-specific gene expression within the stroke subtypes.

**Results:** Our analysis revealed distinct immune cell populations, including monocytes, macrophages, dendritic cells, neutrophils, and T-cells in both clot types. Notably, we observed significant differences in gene expression within the mononuclear phagocytic system cells between clots from atrial fibrillation and carotid atherosclerosis patients.We identified specific genes associated with atherosclerosis and stroke-related processes, such as *CD74, HLA- DRB1*01, HTRA1, C1Q, CD81*, and *CR1* clots from carotid atherosclerosis patients. In atrial fibrillation clots, CD8 T-cells and NK-cells show upregulated expression of genes such as GZMH, GZMB, S100A4, FCGBP2, HLA-A, TIMP1, CLIC1, and IFITM2, indicating their involvement in cytotoxic activities and potential tissue damage. The MAGMA approach highlighted significant genetic associations within leukocytes, particularly in atrial fibrillation and carotid clots, underscoring the potential roles of B-cells, T-cells, and macrophages in clot pathogenesis.

**Conclusions:** This study illuminates the immune and transcriptomic landscape within clots, offering potential biomarkers and lays the foundation for future research.

## INTRODUCTION

Ischemic stroke, a leading cause of mortality and long-term disability, affects millions of individuals annually and requires early intervention for effective treatment.^1^

Previous efforts studying clot composition and its interaction with stroke have provided important findings. For example, studies have investigated how the histological composition of clots is associated with periinterventional clot migration in patients with acute stroke.^3^ These studies have demonstrated the impact of clot composition on technical and clinical success of thrombectomy, as well as its association with worse clinical outcomes.^4^ Additionally, correlations between clot composition and stroke etiology have been explored, revealing differences in clot composition between cardioembolic and large-artery atherosclerosis.^5^ These analyses have shed light on the mechanical properties of clots, which can inform the choice of revascularization strategies and improve thrombectomy outcomes.^6–10^ However, it is important to recognize the limitations inherent to traditional histological studies of clots. These methods, while useful for understanding the macroscopic and structural aspects of clots, often fall short in capturing the complex cellular dynamics and molecular interactions at play. Histological analyses typically provide a static snapshot of clot composition, lacking the depth to elucidate the intricate gene expression patterns and cellular behaviors that drive clot formation and evolution.

In this context, our study aims to investigate the interactions among immune cell types within clots associated with atrial fibrillation (AF), carotid atherosclerosis (CA), employing scRNA-seq to provide a comprehensive understanding of the cellular composition and interactions within these clots.

## METHODS

For a comprehensive description of the methods see **Supplemental Material**.

### Human specimens and patient clinical characteristics

We collected clot samples from 17 patients with large vessel occlusion stroke and deep vein thrombosis undergoing mechanical thrombectomy between October 2020-July 2022 after written informed consent from patients or their surrogates. Mechanical thrombectomy was performed by 2 endovascular surgeons (C.M, R.H) and the differential diagnosis between stroke etiologies was performed by 2 vascular neurologists (D.N, L.S). Eligibility criteria included age ≥18 years, admission to Yale New Haven Hospital, and M1 or M2 occlusion for stroke cases. To protect the identity the subjects, ages are reported as ranges and sex is not reported individually. The present study was overseen by the Yale School of Medicine’s IRB.

### scRNA-seq computational pipelines and analyses

The R package Seurat was used for normalization, data scaling, integration, clustering, dimensionality reduction, differential expression analysis and visualization. Five samples that had mean UMI counts, and mean number of genes detected per cell below 500 were excluded from further analysis. To examine the differences between stroke etiologies, we initially divided the clot samples into two groups: those obtained from patients with confirmed atrial fibrillation (6 samples) and carotid atherosclerosis (4 samples). This allowed us to explore the variations specific to each etiology. Subsequently, to better understand stroke/arterial clots we compared them to venous clots (2 samples), focusing on the distinctions between these types of clots.

### Gene-based and gene-set analyses

We obtained GWAS summary statistics from the GIGASTROKE consortium, encompassing all ischemic strokes, as well as specific subtypes such as cardioembolic stroke and large artery strokes.^6^ **(Supplementary Material)** For control, we also utilized GWAS summary statistics for height and gastroesophageal reflux disease (GERD).^7,8^ The MAGMA approach was employed to assess the association between genetic variants and cell type-specific gene expression within the stroke subtypes.^9^ To complement the GWAS analysis, we integrated single-cell transcriptomic data obtained

## Results

We conducted scRNA-seq on 100,436 cells with an average of 5,908 cells per sample from seventeen clots samples from patients hospitalized with large intracranial vessel occlusion strokes (14) and deep vein thrombosis (3). Following quality control measures, we retained a total of 12 high-quality samples of clots for further analysis. (**Figure 1a)** The demographics and clinical features of these patients are listed in **Table 1**. There were 6 (50%) women among the twelve patients profiled, and the mean age was 74.1 (SD 15.1).

**Figure 1:**
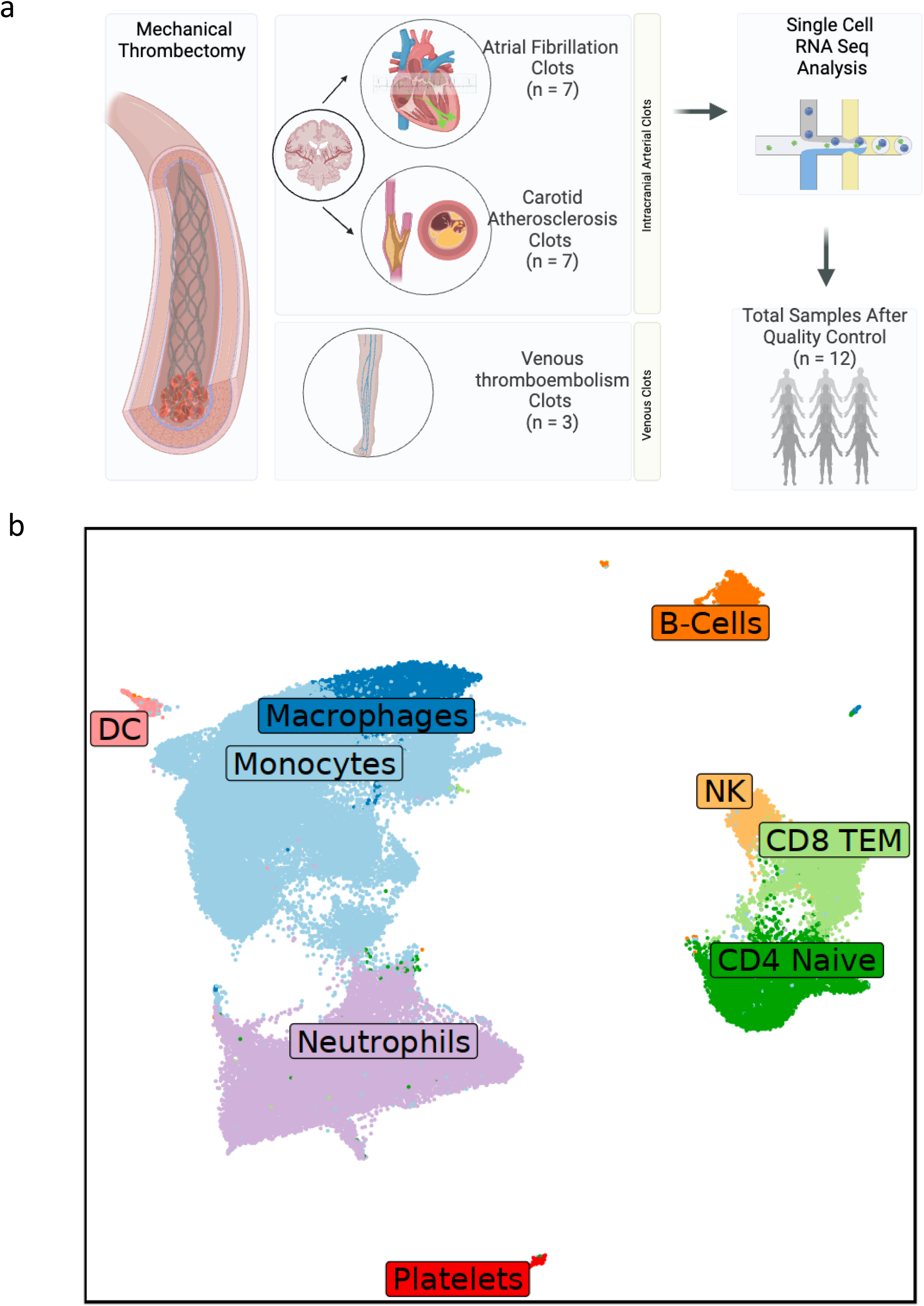
(a) Schematic overview of the study, illustrating the workflow and key objectives. Figure created with BioRender.com. (b) Uniform Manifold Approximation and Projection (UMAP) plot displaying the identified immune cell types within clots.

**Table 1.** Demographics and clinical characteristics of patients.

| Sample | Age | Location of Occlusion | Side | Clinical Diagnosis |
| --- | --- | --- | --- | --- |
| AF1 | 60-69 | M2 | L | Atrial fibrillation |
| AF2 | 70-79 | M1 | L | Atrial fibrillation |
| AF3 | 80-89 | M2 | L | Atrial fibrillation |
| AF4 | 70-79 | M1 | R | Atrial fibrillation |
| AF5 | 80-89 | M1 | R | Atrial fibrillation |
| AF6 | 90-99 | M2 | L | Atrial fibrillation |
| CA1 | 70-79 | M2 | L | Carotid Atherosclerosis |
| CA2 | 80-89 | M1 | L | Carotid Atherosclerosis |
| CA3 | 80-89 | M1-M2 | L | Carotid Atherosclerosis |
| CA4 | 50-59 | ICA | L | Carotid Atherosclerosis |
| VTE1 | 30-39 | Tibioperoneal trunk and left external iliac vein | L | Deep Vein Thrombosis |
| VTE2 | 70-79 | Pulmonary artery bifurcation | - | Pulmonary Embolism |

- \*AF= Atrial fibrillation Clot - \*CA= Carotid atherosclerosis Clot - \*VTE=Venous thromboembolism Clot

### Immune Cell Composition in Thrombotic Clots: A Single-Cell Analysis

Our findings revealed that the predominant immune cell population within the clots consisted mainly of cells belonging to the mononuclear phagocyte system (MPS), particularly monocytes, macrophages, and dendritic cells (DCs). In addition, we observed abundant populations of neutrophils and T-cells within the clots. **(Figure 1b)**

### Transcriptome Analysis of MPS Cells

We found that Macrophages in carotid clots exhibited elevated expression levels of genes involved in antigen presentation (*HLA-DQB1, CD74, and HTRA1*) and immune regulation (*C1QA*, *CD81*, and *CR1*), suggesting their potential role in modulating immune responses and promoting inflammation within the atherosclerotic plaques. **(Figure 2a)** Several studies support these findings, highlighting the involvement of these genes in atherosclerosis and related processes.^10–15^ We observed upregulated expression of various genes in macrophage from atrial fibrillation clots, including genes associated with stress response (*HSPA1B* and *HMOX1*), extracellular matrix remodeling (*TGFBI* and *MMPs*), and immune regulation (*FCER1G* and *CCL2*). **(Figure 2a)** The observed gene expression patterns indicate that macrophages play a role in tissue remodeling and the inflammatory response within atrial fibrillation-associated clots. Notably, *TGFBI* has also been linked to the development of atrial fibrillation.^16^ Gene expression analysis of monocytes in carotid atherosclerosis clots revealed upregulated expression of genes including *FN1, SPP1, FCER1G, MAP3K2, HSP90AA1, SLC25A37, ITM2B*, and *KMT2C*.

**Figure 2:**
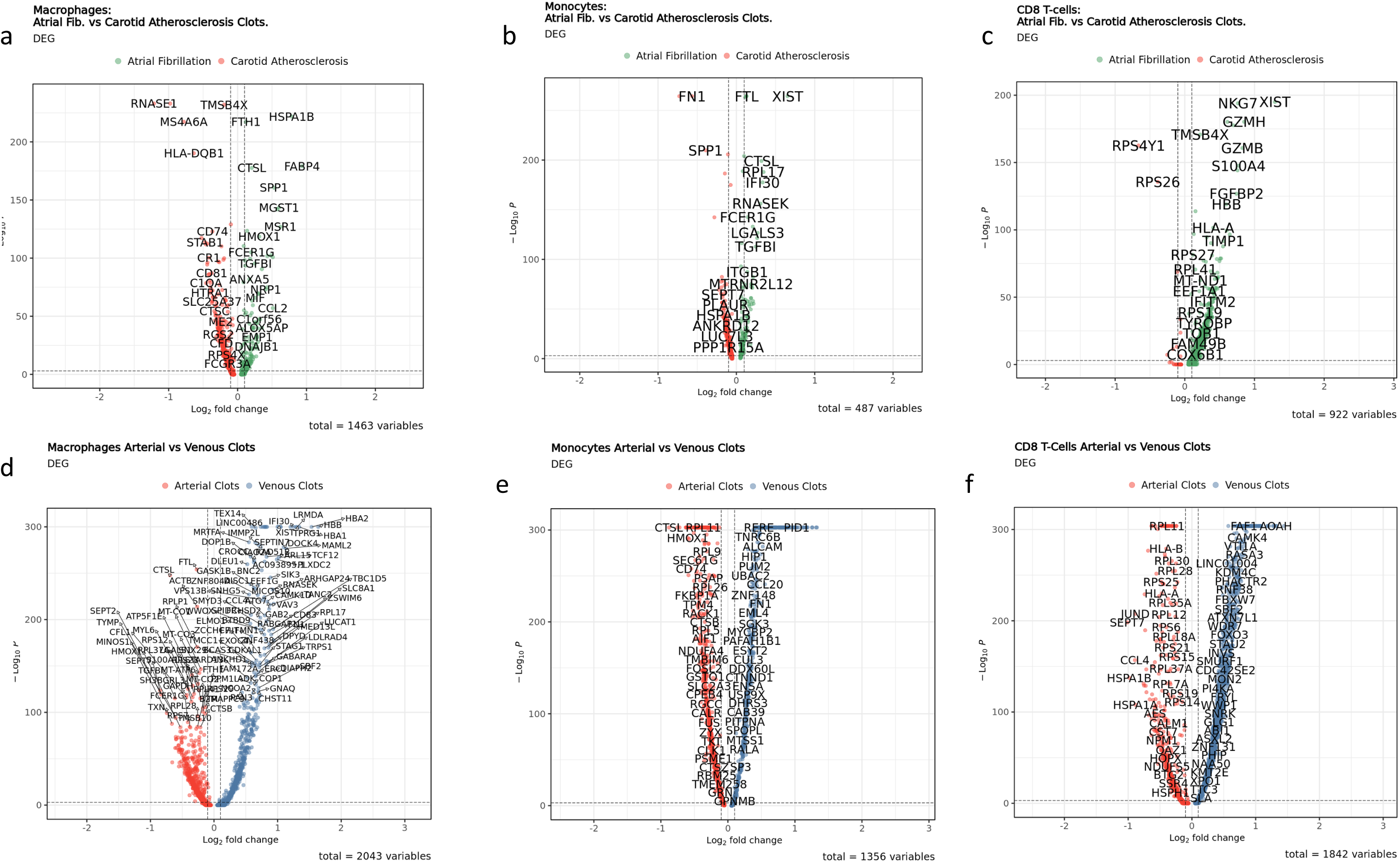
Volcano Plots. (a) Macrophages Differential Gene Expression (DEG) between atrial fibrillation clots and carotid atherosclerosis clots. (b) Monocytes DEG between atrial fibrillation clots and carotid atherosclerosis clots. (c) CD8 T-Cells DEG between atrial fibrillation clots and carotid atherosclerosis clots. (d) Macrophages DEG between venous and arterial clots. (e) Monocytes DEG between venous and arterial clots. (f) CD8 T-Cells DEG between venous and arterial clots.

**(Figure 2b)** Of note, *SPP1* (Osteopontin) plays a significant role in atherosclerosis progression, neointimal hyperplasia, and rapid coronary plaque progression.^17^ Additionally, our findings revealed heightened expression of *FTL*, cathepsin D, and metalloproteinase-9 in monocytes from atrial fibrillation clots, all of which have been extensively discussed in the literature and associated with the pathogenesis of atrial fibrillation.^18–21^ **(Figure 2b)**

Among venous and arterial clots, macrophages from venous clots exhibited increased expression of genes involved in red blood cell breakdown, immune response regulation, calcium homeostasis, RNA degradation, and autophagy (*HBA1, HBA2, HBB, IFI30, LRMDA, MAML2, TPRG1, SLC8A1, RNASEK, CD83, CCL4*, and *GABARAP*). In contrast, macrophages from arterial clots showed increased expression of *FTL, ZNF804A, CTSL, TMCC1, RPS7, MT-ATP6, RPLP1, TYMP, SEPT7, SEPT2, MYL6, TXN, MINOS1*, and *RPL2* genes. These genes participate in various biological processes, including iron storage, transcriptional regulation, protein degradation, cell migration, nucleotide metabolism, cellular organization, muscle contraction, antioxidant activity, and protein synthesis. **(Figure 2d)**

Additionally, monocytes from venous clots exhibited increased expression of *RERE, PID1, TNRC6B, ALCAM, HIP1, PUM2, UBAC2, CCL20, ZNF148, FN1, EML4, SGK3, MYCBP2, PAFAH1B1, ESYT2, CLU3, RALA, MTSS1,* and *SP3*. These genes are implicated in diverse biological functions including cell signaling, adhesion, cytoskeletal organization, immune response, and gene regulation, suggesting a role in venous thrombus formation and stability. In arterial clots, however, monocytes showed heightened expression of *CTSL, RPL11, HMOX1, RRPL9, SEC61G, CD74, PSAP, RPL26, FKBP1A, TPM4, RACK1, CTSB, RPL5, AIF1, TMBIM6*, and *CTSZ*. This gene set is associated with lysosomal activity, protein synthesis, oxidative stress response, antigen presentation, and cell survival, possibly reflecting the distinct mechanical and oxidative stress conditions in arterial thrombosis. **(Figure 2e)**

### Cytotoxicity and Tissue Remodeling: Gene Expression in Atrial Fibrillation Clots

We found that CD8 T-cells and NK-cells from atrial fibrillation clots exhibit upregulated expression of several genes, including *GZMH* (granzyme H), *GZMB* (granzyme B), *S100A4* (S100 calcium-binding protein A4), *FCGBP2* (Fcy-binding protein 2), *HLA-A* (human leukocyte antigen-A), *TIMP1* (tissue inhibitor of metalloproteinase 1), *CLIC1* (chloride intracellular channel protein 1), and *IFITM2* (interferon-induced transmembrane protein 2) **(Figure 2c, Supplementary Material)** The presence of effector markers like *GZMH* and *GZMB* suggests that these lymphocytes are involved in cytotoxic activities and may contribute to tissue damage within the atrial fibrillation clots.

### Ribosomal Biogenesis and Stress Response in CD8 T-cells from Arterial Clot

CD8 T-cells from arterial clots exhibited differential expression of genes associated with ribosomal biogenesis and cellular stress response. Specifically, we observed the upregulation of ribosomal protein genes (RPL genes) and heat shock protein genes, such as *HSPA1B*. These findings suggest the presence of ribosome biogenesis stress within these CD8 T-cells. **(Figure 2f. Supplementary Material)**

### Pathway Expression in Clots

In AF clots, we observed upregulation of many pathways shared across multiple cell types. These included CDC42 signaling, associated with cellular morphology and dynamics, as well as remodeling of epithelial adherens junctions. Our findings also revealed the presence of pathways such as immunogenic cell death signaling, Th1 pathway, RHOA signaling, actin nucleation by ARP-WASP complex, ILK signaling, neutrophil extracellular trap signaling pathway, and hepatic fibrosis signaling pathways were upregulated in AF clots across all cell types. These observations underscore the intricate interplay between immune responses, cellular remodeling, and inflammatory processes in the pathogenesis of AF clots. **(Figure 3a, Supplementary Material)**

**Figure 3:**
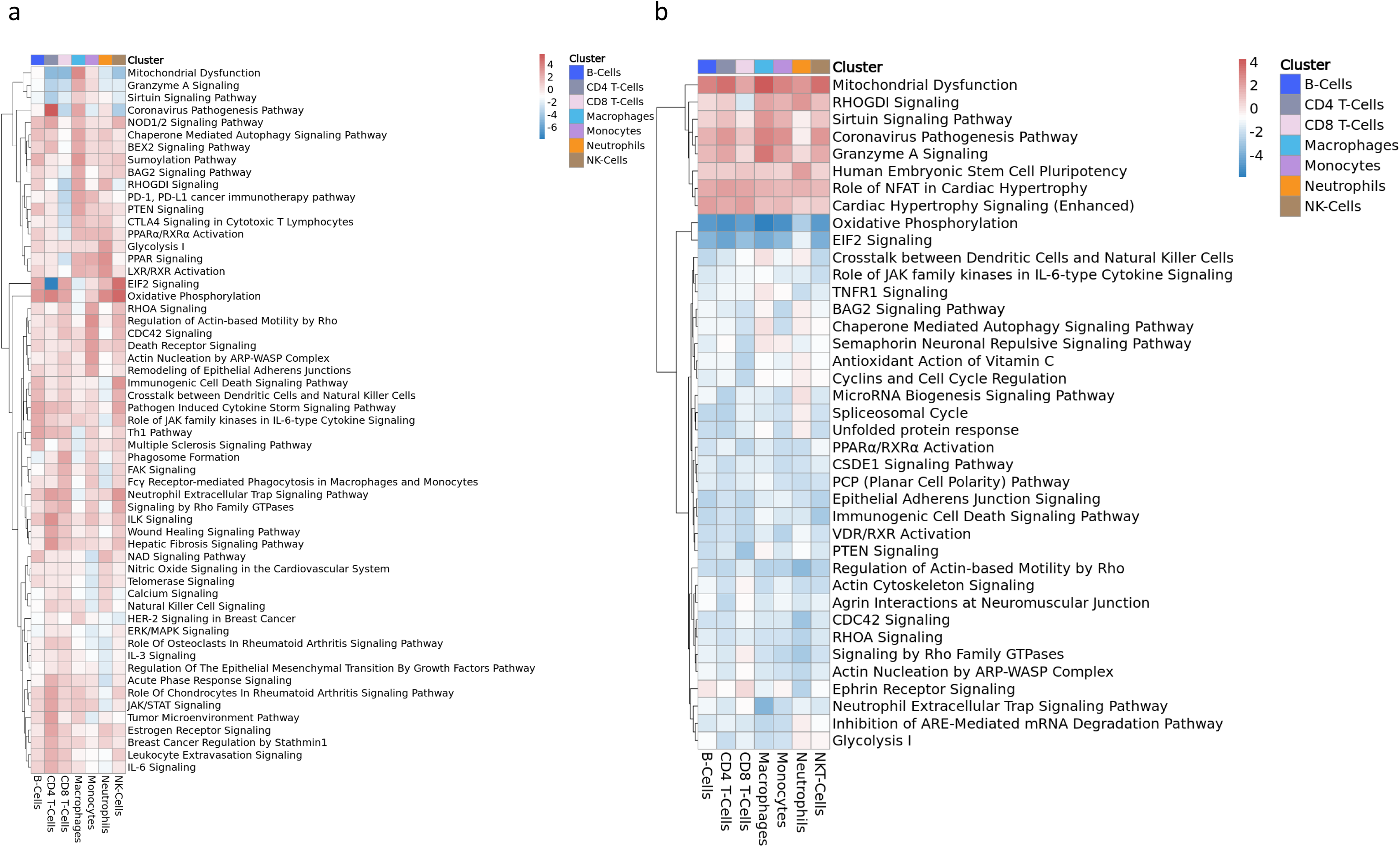
Ingenuity Pathway Analysis (IPA) of Differential Gene Expression (DEG) Results. (a) Heatmap with Ingenuity Pathway Analysis (IPA) of DEG between carotid atherosclerosis clots and atrial fibrillation clots for all clusters (cell types). Red upregulated in FA stroke clots. (b) Heatmap with IPA of DEG between venous and arterial clots. Red upregulated in Venous clots.

In contrast, we found that the specific etiology of carotid atherosclerosis clots led to distinct patterns of pathway expression. Mitochondrial dysfunction, granzyme A signaling, and sirtuin signaling were found downregulated in MPS cells from CA clots. EIF2 signaling, suggesting increased protein translation, was strongly enriched in CD4 T cells from CA clots. In macrophages, Th1 pathway, multiple sclerosis signaling pathway, and phagosome formation were more enriched in the macrophages from CA clots, consistent with activation of macrophages in atherosclerotic lesions. CD8 T cells from CA clots showed upregulation of PD- 1, PD-L1 cancer immunotherapy and CTLA4 signaling suggesting negative regulation of these cells in atherosclerotic clots.

Comparing venous clots to arterial clots, we found upregulated pathways associated with mitochondrial dysfunction, RHODI signaling, Sirtuin signaling pathway, Coronavirus pathogenesis pathways, Granzyme A signaling, Human Embryonic Stem Cell pluripotency, and Role of NFAT in cardiac hypertrophy and cardiac hypertrophy signaling in venous clots. **(Figure 3b, Supplementary Material)**. Conversely, in arterial clots, we observed upregulated pathways related to oxidative phosphorylation, EIF2 signaling, crosstalk between DC and natural killers, Role of JAK family kinases in IL-6 type cytokine signaling, TNFR1 signaling, Epithelial Adhesion Junction signaling, Inflammatory Cell Death signaling pathway, PTEN signaling, VDR/RXR activation, Actin Cytoskeleton signaling, CDC42 signaling, RHOA signaling, and Neutrophil Extracellular Trap signaling pathway. These findings reveal potential alterations in cellular energetics, protein synthesis, immune cell communication, cytokine signaling, cell adhesion, inflammatory responses, and cytoskeletal dynamics within arterial clots.

### Receptor-Ligand Insights

We investigated the receptor-ligand interactions within various cell populations. In AF clots, we observed a higher number of significant interactions involving DC compared to other cell types. **(Figure 4a)** Notably, significant interactions were found between DC and DC, DC and monocytes, and DC and platelets. In carotid atherosclerosis clots, we found a higher prevalence of significant interactions involving natural killer (NK) cells and neutrophils. Specifically, significant interactions were observed between NK cells and NK cells, NK cells and neutrophils, and neutrophils and neutrophils NK. **(Figure 4b)** Venous clots exhibited a distinct pattern of significant interactions, with a notable emphasis on interactions involving DC. The most prominent interactions observed were DC-DC interactions, as well as interactions between DC and neutrophils, NK cells, and platelets. **(Figure 4c)** Additionally, we investigated the presence of unique interactions among these cells in each stroke type. In atrial fibrillation, we identified specific interactions among DC-DC that included VCAM1-ITGA4/ITGB7, JAG1- NOTCH2/NOTCH4, and CD274-PDCD1. These interactions hold biological significance, as they implicate molecules involved in cell adhesion, immune signaling, and immune checkpoint regulation. Furthermore, in carotid clots, the unique interactions among DC-DC encompassed intriguing pairs such as TGFB1-ACVR1, VEGFA-FTL1, IL10-IL10RA, CXCL1-CX3CR1, and ADGREES-CD55/LPAR2/LPAR3. These interactions suggest potential involvement of these molecules in modulating inflammatory responses, angiogenesis, and cellular signaling pathways specifically within the context of carotid atherosclerosis-related thrombosis. **(Figure 4d-e)**

**Figure 4:**
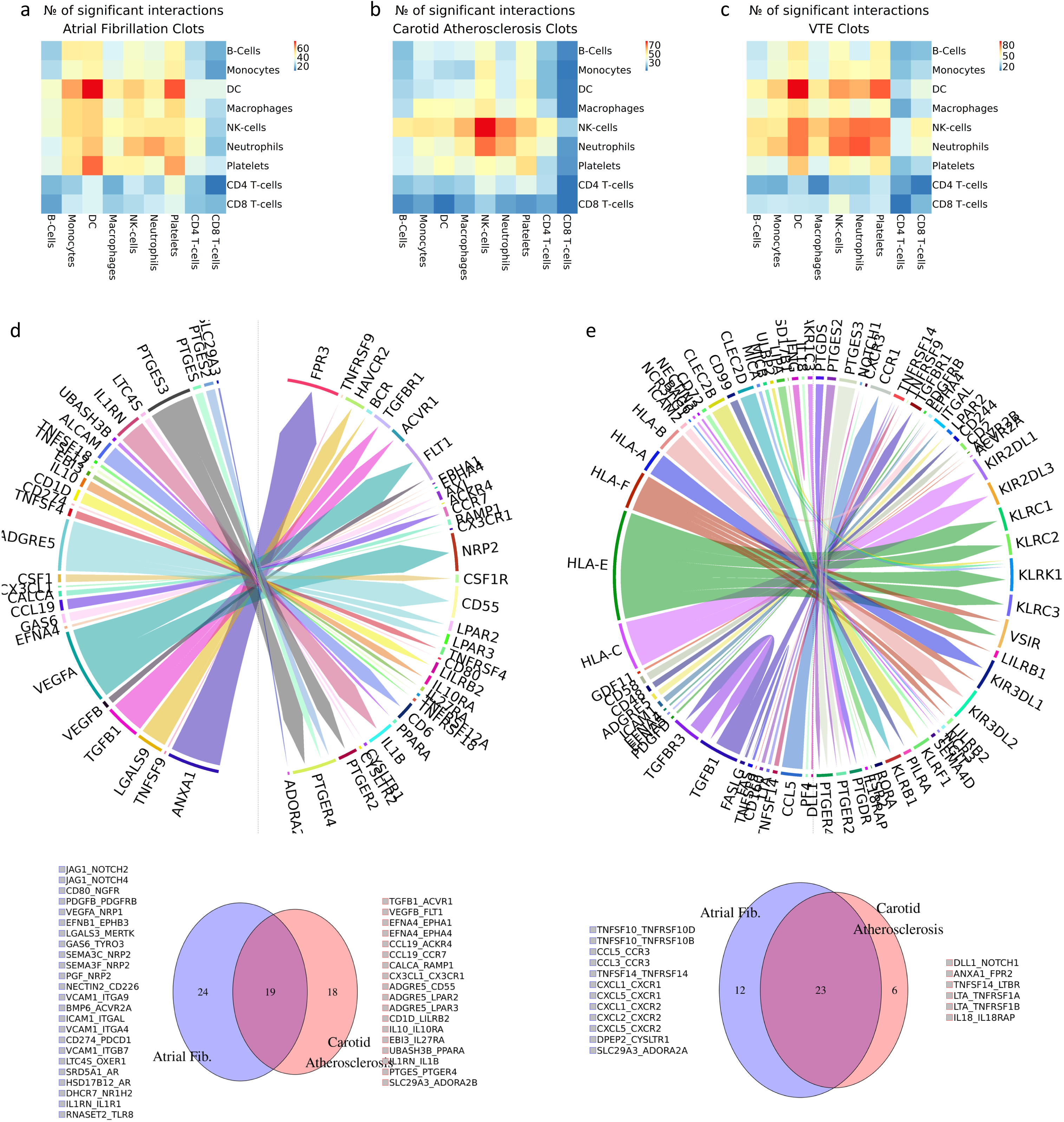
Interactions Among Immune Cell Types in Atrial Fibrillation and Carotid Atherosclerosis Clots. (a) Heatmaps displaying the number of significant interactions among cell types in atrial fibrillation (AF) clots. (b) Heatmaps displaying the number of significant interactions among cell types in Carotid Atherosclerosis clots. (c) Heatmaps displaying the number of significant interactions among cell types in Venous Clots. (d) Circos plot illustrating significant interactions between dendritic cells (DC) within AF clots and VennDiagram showing the number of significant interactions shared and the number of unique interactions between etiologies in DC cells. (e) Circos plot illustrating significant interactions between natural killer cells (NK-Cells) within AF clots and VennDiagram showing the number of significant interactions shared and the number of unique interactions between etiologies in NK-cells. \**The legend accompanying the Venn diagrams provides the names of specific unique interactions*.

### Genetic Insights into Clot Formation in Atrial Fibrillation and Carotid Clots

We examine the expression of known genetic risk variants for stroke in leukocytes from the different clot types. Our comprehensive analysis utilizing the GIGASTROKE trans-ancestry GWAS dataset uncovered significant associations within the genetic landscape of clot formation in atrial fibrillation and carotid clots. **(Figure 5, Supplementary Material)** By employing MAGMA, we ranked gene expression specificity for each cell type and observed intriguing patterns in the context of clot pathogenesis. **(Figure 5a)** In atrial clots, B-cells, CD4 T-cells, CD8 T-cells, macrophages, monocytes, and DC surpassed the predetermined p-value threshold, indicating their potential roles in the intricate mechanisms of stroke development. **(Figure 5b)** Similarly, in carotid clots, B-cells, CD8 T-cells, CD4 T-cells, NK cells, and macrophages exhibited significant associations, underscoring their relevance in the genetic factors governing carotid clot formation. **(Figure 5c)** As controls, we used gene sets for gastrointestinal reflux and height and failed to find significant enrichment in the leukocytes from the clots. These findings shed light on the active participation of these specific cell types in particular T-cells and their interactions in the complex molecular processes underlying stroke.

**Figure 5:**
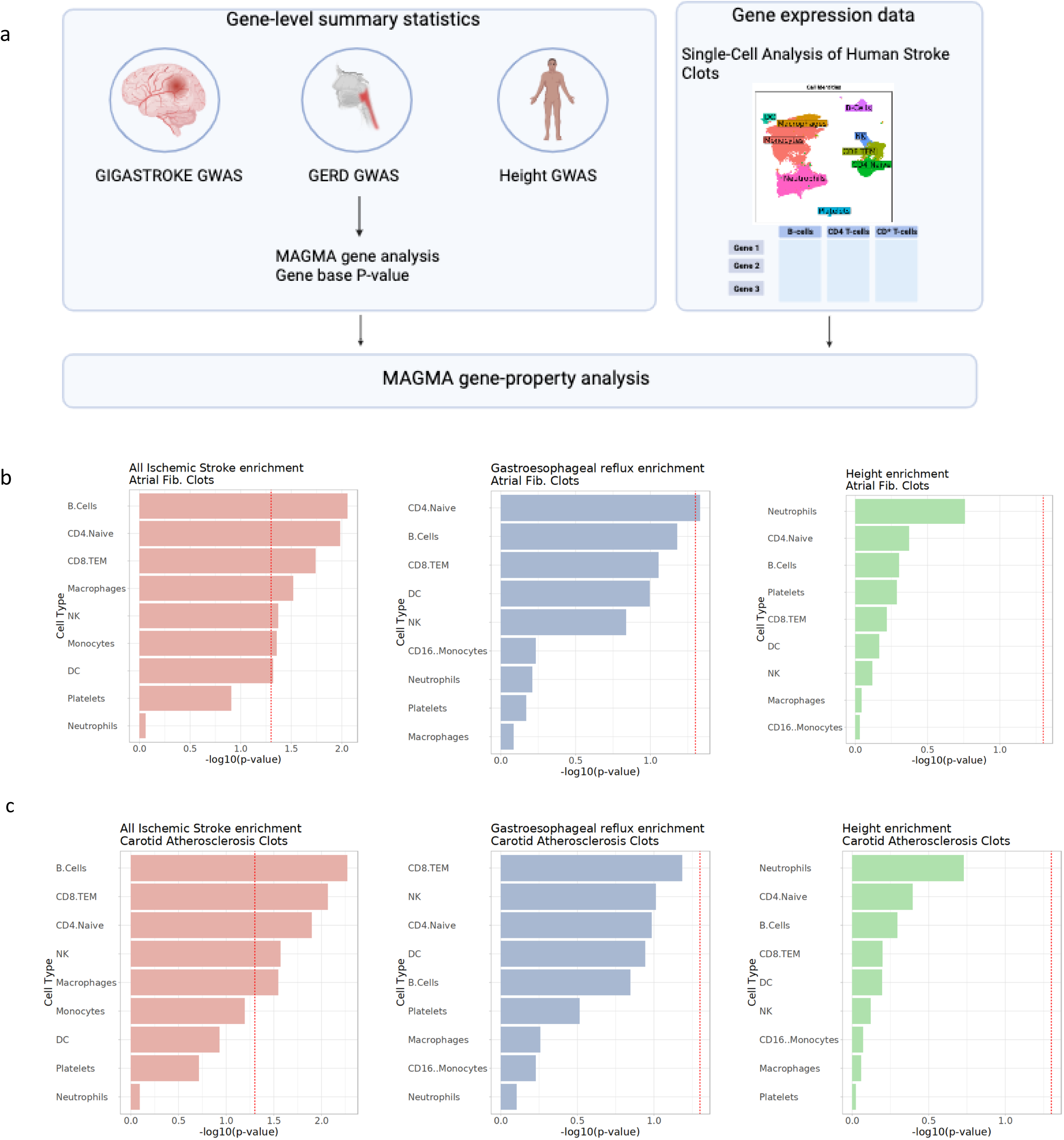
MAGMA Workflow and Cell-Type Analysis in Human Stroke Clots. (a) Schematic representation of the MAGMA workflow utilizing the GIGASTROKE, Gastroesophageal reflux disease (GERD) and height GWAS. (b) Bar plots depict the relationship between various cell types in Atrial Fib. Clots and the p-values obtained from the MAGMA gene analysis for each different GWAS. The red lines represent the p-value threshold of <0.05, indicating statistically significant associations. (c) Bar plots depict the relationship between various cell types in Carotid Atherosclerosis Clots and the p-values obtained from the MAGMA gene analysis for each different GWAS. The red lines represent the p-value threshold of <0.05, indicating statistically significant associations.

## Discussion

In our study, we employed scRNA-seq analysis to investigate the cellular composition, transcriptome and interactions within stroke clots. Our analysis revealed the predominant presence of immune cells belonging to the MPS within the clots, including monocytes, macrophages, and dendritic cells. Neutrophils and T-cells were also observed in abundance. Interestingly, we found variations in the composition of immune cell populations between different etiologies of stroke (cardioembolic versus carotid disease). Carotid atherosclerosis clots showed enrichment of B-cells, CD4 T-cells, dendritic cells, and macrophages, while atrial fibrillation clots exhibited enrichment of CD8 T-cells, monocytes, NK cells, and neutrophils. This suggests distinct immune responses and mechanisms associated with different etiologies.

Furthermore, the analysis of gene expression patterns provided insights into potential stroke biomarkers. We identified specific genes associated with atherosclerosis and stroke-related processes, such as *CD74, HLA-DRB1*01, HTRA1, C1Q, CD81*, and *CR1*. In addition, we observed differential gene expression patterns in CD8 T-cells and NK cells from atrial fibrillation clots, indicating their involvement in cytotoxicity, tissue remodeling, and antigen presentation.

Comparative analysis of arterial and venous clots revealed distinct gene expression profiles in macrophages, suggesting unique molecular characteristics and potential roles in clot formation and associated pathophysiological processes. Additionally, pathway analysis unveiled dysregulated pathways associated with mitochondrial dysfunction, immune response, cellular stress response, and inflammatory processes in both atrial fibrillation and carotid atherosclerosis clots. Moreover, our study explored cell-cell interactions within the clot, this analysis enabled us to identify potential communication networks and interactions between immune cells and other cell types present in the clots.

It is important to note that while our study provides valuable insights, there are limitations to consider. The sample size was relatively small, and further validation with larger cohorts is necessary to confirm the observed findings. Moreover, the analysis focused on scRNA-seq data, and functional validation of the identified biomarkers and pathways is required.

In summary, our study highlights the utility of scRNA-seq in characterizing the immune landscape of clots associated with stroke. The findings shed light on the cellular composition, gene expression profiles, and pathways involved in different etiologies, providing insights into potential biomarkers and therapeutic targets. Further research and validation are needed to translate these findings into clinical applications that can improve the diagnosis, treatment, and management of stroke and venous thrombosis.

## Abbreviations

AF: Atrial Fibrillation
CA: Carotid Atherosclerosis DC - Dendritic Cells
MPS: Mononuclear Phagocyte System NK –
Natural Killer scRNA-seq: Single-cell RNA Sequencing
UMAP: Uniform Manifold Approximation and Projection

## Code availability

All data and scripts used for data analysis are available at GitHub (https://github.com/dadarenedo/human_clots).

## Authors Contribution

Conceptualization (C), Methodology (M), Formal Analysis (FA), Data Curation (DC), Writing - Original Draft (WOD), Writing - Review & Editing (WRE), Visualization (Vis), Figure Preparation (FP), Supervision (Sup), Project Administration (PA), Funding Acquisition (F).

**DR:** C, M, FA, DC, WOD, Vis, Sup, FP, PA.

**TB:** C, M, S, FA, DC, WOD.

**JD:** M.

**JNA:** C, M, FA, Vis, FP.

**NS:** WRE. **JA:** WRE. **AK:** WRE.

**CR:** FA, WRE.

**SCT:** FA, WRE.

**SH:** FA, WRE.

**JG:** WRE.

**GJF:** WRE.

**KNS:** WRE.

**RH:** WRE.

**MG:** WRE.

**LHS:** C, M, WRE, Sup, PA, F.

**DSN:** C, M, WRE, Sup, PA, F.

**CM:** C, M, WRE, Sup, PA, F.

## Notes

### Competing Interest Statement

The authors have declared no competing interest.

### Funding Statement

no external funding was received

### Author Declarations

The present study was overseen by the Yale School of Medicine's IRB.

